# Nitrogen dioxide exposures from LPG stoves in a cleaner-cooking intervention trial

**DOI:** 10.1101/2020.06.25.20139345

**Authors:** Josiah L Kephart, Magdalena Fandiño-Del-Rio, Kendra N Williams, Gary Malpartida, Alexander Lee, Kyle Steenland, Luke P. Naeher, Gustavo F. Gonzales, Marilu Chiang, William Checkley, Kirsten Koehler, CHAP trial Investigators

## Abstract

**Background:** Liquefied petroleum gas (LPG) stoves have been promoted in low- and middle-income countries (LMICs) as a clean energy alternative to biomass burning cookstoves.

**Objective:** We sought to characterize kitchen area concentrations and personal exposures to nitrogen dioxide (NO_2_) within a randomized controlled trial in the Peruvian Andes. The intervention included the provision of an LPG stove and continuous fuel distribution with behavioral messaging to maximize compliance.

**Methods:** We measured 48-hour kitchen area NO_2_ concentrations at high temporal resolution in homes of 50 intervention participants and 50 control participants longitudinally within a biomass-to-LPG intervention trial. We also collected 48-hour mean personal exposures to NO_2_ among a subsample of 16 intervention and 9 control participants. We monitored LPG and biomass stove use continuously throughout the trial.

**Results:** In 367 post-intervention 24-hour kitchen area samples of 96 participants’ homes, geometric mean (GM) highest hourly NO_2_ concentration was 138 ppb (geometric standard deviation [GSD] 2.1) in the LPG intervention group and 450 ppb (GSD 3.1) in the biomass control group. Post-intervention 24-hour mean NO_2_ concentrations were a GM of 43 ppb (GSD 1.7) in the intervention group and 77 ppb (GSD 2.0) in the control group. Kitchen area NO_2_ concentrations exceeded the WHO indoor hourly guideline an average of 1.3 hours per day among LPG intervention participants. GM 48-hour personal exposure to NO_2_ was 5 ppb (GSD 2.4) among 35 48-hour samples of 16 participants in the intervention group and 16 ppb (GSD 2.3) among 21 samples of 9 participants in the control group.

**Discussion:** In a biomass-to-LPG intervention trial in Peru, kitchen area NO_2_ concentrations were substantially lower within the LPG intervention group compared to the biomass-using control group. However, within the LPG intervention group, 69% of 24-hour kitchen area samples exceeded WHO indoor annual guidelines and 47% of samples exceeded WHO indoor hourly guidelines. Forty-eight-hour NO_2_ personal exposure was below WHO indoor annual guidelines for most participants in the LPG intervention group, and we did not measure personal exposure at high temporal resolution to assess exposure to cooking-related indoor concentration spikes. Further research is warranted to understand the potential health risks of LPG-related NO_2_ emissions and inform current campaigns which promote LPG as a clean-cooking option.

## 1. Introduction

Nearly 40% of the global population uses biomass fuels as their primary source of energy for cooking.^1^ Biomass cookstove emissions often result in high levels of household air pollution (HAP), a leading environmental contributor to the global burden of disease and the cause of an estimated 1.6 million premature deaths in 2017.^2^ Exposure to HAP has been associated with increased blood pressure,^3,4^ lung cancer,^5,6^ and COPD ^7–10^ in adults. Women and their children are particularly vulnerable to biomass smoke exposure due to their proximity to cooking activities in many settings.^11^ The existing HAP literature has focused on fine particulate matter (PM_2.5_) and carbon monoxide (CO) as the components of biomass emissions which are most relevant to public health.^2,12^ However, nitrogen dioxide (NO_2_), an air pollutant causally related to poor respiratory outcomes,^13^ has also been reported in homes with biomass cookstoves at concentrations which exceed WHO indoor air quality guidelines.^14–23^

To reduce HAP exposures and prevent HAP-related disease, most public health interventions have focused on improved biomass cookstoves, which aim to reduce HAP exposures by improving stove combustion efficiency and/or directing stove emissions outdoors, often while continuing to rely on locally available biomass fuels.^24^ Although emissions from these improved cookstoves are often lower than traditional cookstoves, concentrations and exposures from improved biomass cookstoves generally remain above WHO indoor guidelines.^25,26^ More recently, international campaigns ^27^ and national governments ^28,29^ have promoted liquefied petroleum gas (LPG) as a cleaner-burning alternative to biomass fuels. LPG is typically transported in portable cylinders that are connected to a stove by a hose. LPG is becoming a common household fuel in many urban areas of low- and middle-income countries (LMICs).^30^

These LPG stoves appear to be effective at reducing emissions of PM_2.5_ and CO ^26,31–35^ to levels which could provide substantial public health benefits.^36^ However, a recent study of nearly 76,000 gas and electricity users in China found lower all-cause mortality in homes with vs. without kitchen ventilation,^37^ suggesting that even “clean” fuels can produce health-altering emissions. Beyond PM_2.5_ and CO, little is known about the effect of transitioning from biomass to LPG stoves on other household air pollutants, including NO_2_.

NO_2_ is a widely regulated ambient air pollutant ^38,39^ that is considered by the United States Environmental Protection Agency (US EPA) to be causally related to respiratory effects.^13^ The most established health effects associated with NO_2_ include pediatric asthma^40,41^ and reduced lung function.^42–49^ A growing body of literature suggests associations between NO_2_ exposure and cardiovascular, respiratory, and all-cause mortality.^50–52^. In high income countries (HICs), natural gas is a common household fuel, and natural gas-burning appliances such as stoves, ovens, and heaters can be significant household sources of indoor NO_2_.^13,53–55^ NO_2_ concentrations in homes with gas appliances in HICs can often meet or exceed WHO indoor annual guidelines,^53–57^ and indoor NO_2_ concentrations in such homes have specifically been associated with respiratory symptoms in children.^53^ Stove quality, maintenance, design, and gas fuel type (i.e. natural gas, LPG) are known to impact emissions of NO_2_ from gas stoves.^26,58^ However, nearly all assessments of NO_2_ exposure from gas appliances have taken place in HICs. Given the known elevated concentrations of indoor NO_2_ from natural gas stoves in HICs and the plausible differences between primary fuel type, gas stove design, function, and quality between HICs and LMICs, there is a need for direct measurement of NO_2_ exposures from LPG stoves in LMIC settings. This information is critical to inform the promotion of LPG stoves as an effective public health intervention. This study aims to characterize the impact of a biomass-to-LPG intervention trial on kitchen area concentrations and personal exposures to NO_2_ in the Peruvian Andes. As a secondary analysis to inform HAP exposure assessment strategies, we analyzed between-participant versus within-participant variance across 1) two consecutive 24-hour samples and 2) two 24-hour samples taken months apart during the post-intervention period.

## 2. Methods

### 2.1. Study design and setting

We conducted a randomized controlled trial of a cleaner-cooking intervention among women who used biomass cookstoves in the Peruvian Andes. The study took place in the Puno region of southern Peru, bordering Lake Titicaca and located approximately 3,825 meters above sea level. Puno is a rural agricultural region where subsistence farming, alpaca husbandry, and small-scale quinoa and potato production are common. Study participants were enrolled from Indigenous Aymara communities where Spanish and Aymara are commonly spoken. In these low-density communities, homes are a median distance of 101 meters from the closest neighboring house.^59^ Local sources of ambient air pollution are minimal and only 4% of houses in the study area are within 100 meters of an arterial road.^59^

In the Cardiopulmonary outcomes and Household Air Pollution (CHAP) trial,^59^ 181 women between the ages of 25 – 64 years were enrolled and randomized 1:1 into an LPG intervention arm and a control arm. One control participant withdrew from the study after baseline assessments, leaving an intention-to-treat sample of 180 participants. Participants in the LPG intervention arm received a free, three-burner LPG cookstove (**Figure 1**) installed by trained research staff, free LPG fuel delivered as needed for one year, as well as education and behavioral reinforcement of exclusive LPG stove use. Participants in the control arm continued to use biomass and will receive a free LPG stove and one-year of fuel the following year.

**Figure 1.**
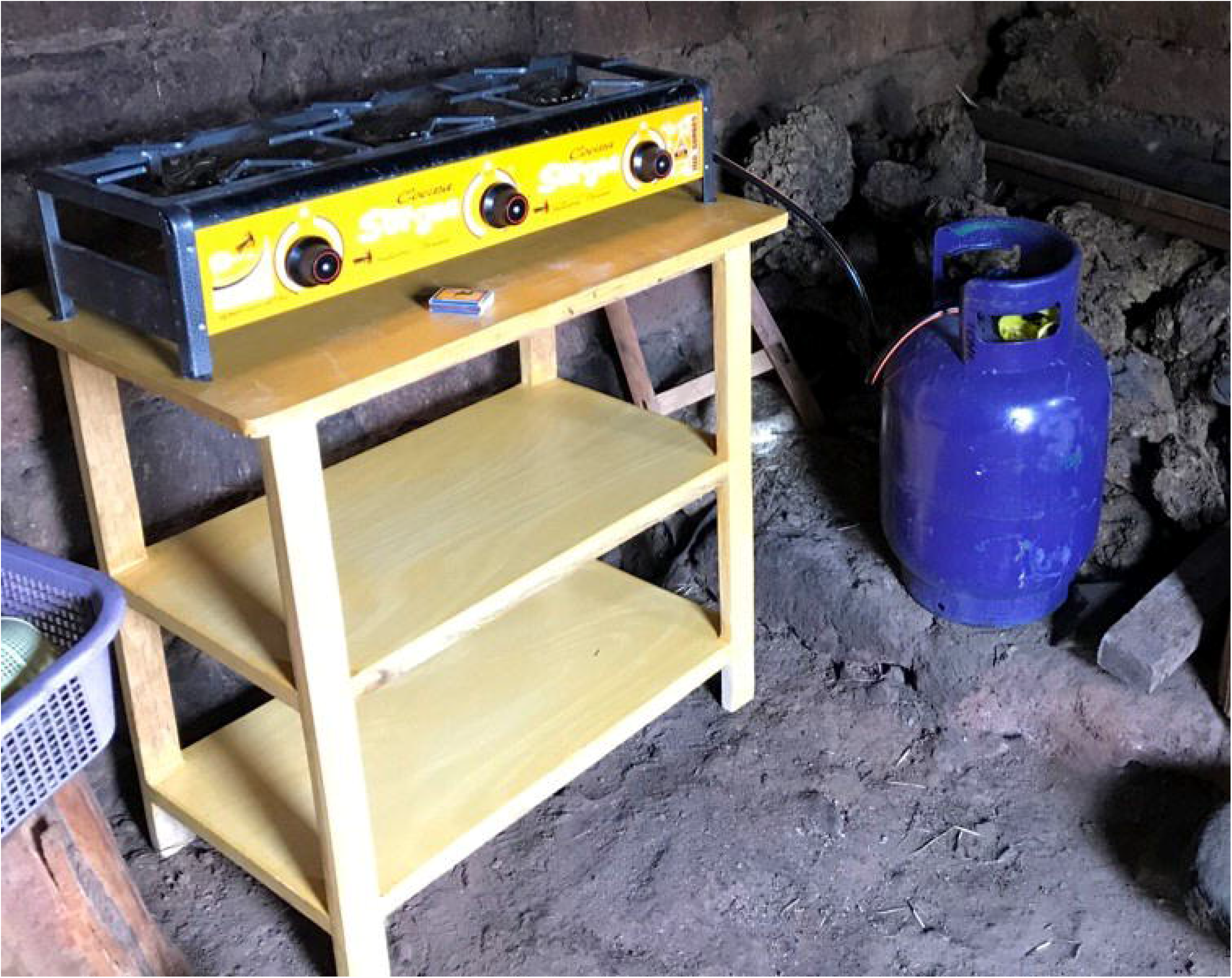
Three-burner LPG stove with table and LPG cylinder, as installed in the kitchens of participants in the intervention group of an LPG cleaner-cooking trial in Puno, Peru.

Eligibility criteria included daily use of biomass fuels for cooking, full-time residence in their current location for at least six months, and being the primary cook for the household. Women were excluded if they had hypertension, COPD, or pulmonary tuberculosis, smoked cigarettes daily, were pregnant or planned to become pregnant within the next year, or if they planned to move out of the study area in the coming year. Demographic information was collected at baseline via questionnaires and HAP assessments were performed at baseline and at three, six, and 12 months post-intervention. NO_2_ exposure in 100 homes with biomass cookstoves using the CHAP trial baseline assessments ^14^ and further information on the CHAP trial study design and assessments has been previously published.^59^

We sampled kitchen area NO_2_ concentrations during the CHAP trial’s post-intervention, follow-up period in a randomized subsample of 100 participants from the larger trial (n = 180). All subsequent references to intervention and control groups refer to this subset of 100 participants. Of the subset of 100 participants, 25 participants were randomly selected for additional assessment of personal exposure to NO_2_. All participants gave verbal informed consent and study protocols were approved by the Johns Hopkins School of Public Health Institutional Review Board (00007128), A.B. PRISMA Ethical Institutional Committee (CE2402.16), and the Universidad Peruana Cayetano Heredia Institutional Review Board (66780).

### 2.2. Nitrogen dioxide exposure assessment

#### 2.2.1. Kitchen area assessment

NO_2_ kitchen area concentrations were measured at one-minute resolution with direct-reading instruments for a targeted 48 hours at baseline ^14^ and three, six, and 12 months post-intervention. A randomly selected subsample of 25 kitchens was also assessed using passive time-integrated samplers. Direct-reading and passive samplers (when applicable) were co-located in wire bird cages and hung from the ceiling of participants’ kitchens. Trained research staff used measuring tapes to place monitors 1.5 meters above the floor and 1.0 meter horizontally from the edge of the cookstove combustion zone, avoiding windows and doors as much as possible, to approximate the breathing zone of a woman tending the fire.

To measure kitchen area NO_2_ concentrations at high-temporal resolution, we used Aeroqual Series 500 portable monitors with NO_2_ sensor heads (Aeroqual Limited, Auckland, New Zealand). These direct-reading monitors were supported by two auxiliary batteries due to limited electricity in participant homes. Every four months, we co-located all direct-reading monitors in the field office to assess imprecision between devices. Using an LPG stove as a source of NO_2_ emissions, we subjected all co-located monitors to NO_2_ concentrations ranging continuously from background concentration to approximately 1000 ppb and back to background concentration. We then estimated the median measurement from all co-located sensors at each minute of the colocation. We used robust linear regression with Siegel repeated medians (*mblm* R package v0.12.1; Komsta, 2019) to calculate intercept and slope adjustments for each sensor, adjusting each sensor to the median concentration observed across the continuous range of concentrations (background to approximately 1000 ppb). To determine the limit of detection (LOD), two direct-reading monitors were brought to the Johns Hopkins University in Baltimore, USA for co-location with a gold-standard reference instrument (model 42c, Thermo Environmental Instruments Inc., Franklin, MA, USA). We defined the LOD as three times the standard deviation (SD) of the difference between the device measurements and reference instrument-confirmed zero-air from a dynamic gas calibrator (model 146i, Thermo Environmental Instruments Inc., Franklin, MA, USA). We estimated an LOD of 20 ppb for the direct-reading monitors, and 35% of all collected 1-minute measurements during the post-intervention period fell beneath the LOD. All concentrations < 20 ppb were replaced with LOD/sqrt(2) ≈ 14.1 ppb, which is similar to a recent modeled estimate of annual ambient NO_2_ concentrations in the Puno region (≈ 12 ppb).^60^ We decommissioned NO_2_ sensor heads after twelve months of field sampling and replaced with new, factory-calibrated sensor heads, as recommended by the manufacturer for high concentration settings.

We sampled time-integrated kitchen area NO_2_ concentrations in a subset of 25 households using Ogawa passive samplers (Ogawa USA, Pompano Beach, FL, USA). We used standard colorimetric methods ^61^ to analyze the passive samples at the Universidad Peruana Cayetano Heredia in Lima, Peru. We measured temperature and relative humidity during each sample with a collocated Enhanced Children’s Monitor (RTI Inc., Research Triangle Park, NC, USA) ^62^ to assist in calculating final NO_2_ concentrations. Temperature data for one sample was missing due to instrument failure and was imputed using the median temperature from all kitchen samples. We took passive sampler field blanks every 10^th^ sample and calculated the LOD as the mean plus SD*3 concentration among blanks. We estimated an LOD of 2.6 ppb, similar to the manufacturer recommended lower range of accuracy (2 ppb). One of the passive sampler kitchen area concentrations (4%) fell beneath the LOD and was replaced with LOD/sqrt(2) ≈ 1.8 ppb.

#### 2.2.2. Personal exposure assessment

We assessed personal exposure to NO_2_ for 48 hours among 25 participants using Ogawa passive samplers as described previously. These badge samplers are small, lightweight, and can be easily worn by participants, in contrast to the direct-reading monitors used for kitchen area sampling which allow for measurements at high temporal resolution but are bulkier and heavier. We altered aprons that are commonly worn by women in the study setting and attached the NO_2_ sampler and temperature and humidity monitors to the central chest region, to approximate each woman’s breathing zone. Field staff demonstrated how to put on and remove the device-laden aprons, and requested that participants wear the aprons at all times during waking-hours and place the apron nearby when bathing or sleeping. Two personal samples had missing temperature data, which were replaced with the median temperature among all personal samples. We used the same passive sampler LOD of 2.6 ppb for personal exposure samples, and we replaced seven personal exposure samples (18%) that were below the LOD with LOD/sqrt(2) ≈ 1.8 ppb.

### 2.3. Stove use monitoring

The temperature of each LPG stove was monitored every minute throughout the duration of the study using Digit-TL temperature loggers with aluminum encasings (LabJack Corporation, Lakewood, CO, USA). As higher stove temperatures indicate cookstove use, temperature loggers have become an important method of directly monitoring stove use in cookstove studies, ^63–66^ commonly referred to as Stove Use Monitors (SUMs). We suspended a temperature logger from the middle burner of each LPG stove. To monitor biomass cookstoves, we attached temperature loggers as close as possible to the cooking surface of the cookstove, typically within the smoke stream and within 1.0 meters of the combustion zone. Additional information on the SUMs methods are included as a supplement.

### 2.4 Statistical Methods

#### 2.4.1 Analysis of nitrogen dioxide measurements

We hypothesized that mean kitchen area concentrations were highly driven by short-term concentration spikes associated with a small number of cooking events per day. To avoid bias from variability in the duration of samples (and the number of cooking events contained in that duration), we calculated 24-hour mean concentrations for each of the two days if at least 20 hours of measurement data was available. Due to battery failure, 66 of 352 total direct-reading samples (19%) had durations < 20 hours and were excluded from the analysis. For 64 of 352 total samples (18%) with durations between 20 and 44 hours, we used the first 24 hours to calculate 24-hour means (of which three samples had durations between 20 – 24 hours and the full available duration was considered a 24-hour mean). A total of 222 of 352 samples (63%) lasted >= 44 hours and two 24-hour mean concentrations were calculated (1^st^ day and 2^nd^ day of total sample).

Because of the high-altitude setting in Puno, we assumed an altitude of 3825 MASL and conditions of 10°C to estimate an atmospheric pressure of 625 hPa and convert mass concentration WHO indoor guidelines to conditions-adjusted ppb (annual, 40 µg/m^3^ = 33 ppb; hourly 200 µg/m^3^ = 163 ppb).^23^ We calculated hourly mean concentrations as the centered, rolling 60-minute mean during each 24-hour sample. We also calculated the proportion of time in which kitchen concentrations exceeded 163 ppb, the conditions-adjusted WHO indoor hourly guideline,^23^ and derived the number of daily hours in excess of the indoor hourly guideline. We calculated summary statistics for the maximum hourly mean, 24-hour mean, and daily hours in excess of 163 ppb. Using the SUMs results and the direct-reading monitors, we calculated mean kitchen area NO_2_ concentrations during cooking events and outside of recorded cooking events. Finally, we calculated summary statistics for the time-integrated passive badge samples of kitchen area concentration and personal exposure.

#### 2.4.2 Stove use analysis

We developed separate empirical algorithms to predict LPG and biomass cookstove use with recorded stove temperatures from the SUMs. Additional information on statistical methods is included as a supplement.

#### 2.4.3. Analysis of effect of LPG stove intervention on NO_2_ concentrations

To assess longitudinal changes in NO_2_ concentrations over the course of the 12-month post-intervention period, we used a one-way ANOVA to examine marginal differences in mean kitchen area concentrations between post-intervention time points within the LPG intervention and control households separately.

In baseline measurements,^14^ we observed differences in kitchen area mean NO_2_ concentration between treatment groups despite randomization (one-way ANOVA, mean NO_2_ concentrations 32 ppb lower in LPG intervention group than control group at baseline, p = 0.04, N = 143 24-hour means). To assess whether differences in post-intervention NO_2_ concentrations were associated with the intervention versus the result of baseline differences between treatment groups, we used linear regression to estimate the effect of the intervention on kitchen area NO_2_ concentrations during the entire post-intervention period, adjusting for baseline concentrations. We used a single time-weighted-average (TWA) concentration for each household at each post-intervention time point for this longitudinal analysis, averaging the 1^st^ and 2^nd^ day 24-hour means from each sample when available (N=160 48-hour samples) and using the 1^st^ day 24-hour mean if the sample did not last long enough to provide a valid 2^nd^ day 24-hour mean (N=47 24-hour samples).

#### 2.4.4. Analysis of variance of 1^st^ versus 2^nd^ consecutive sampling days

We analyzed the reproducibility of 24-hour sampling by comparing consecutive 1^st^ and 2^nd^ day mean kitchen area concentrations among all post-intervention samples that achieved two full days of sampling (44 - 48 hours total duration). We observed heteroscedasticity in the residuals which violated model assumptions and was resolved by log-transforming NO_2_ concentrations for the final analysis. We performed a one-way mixed effects ANOVA assessing between-participant and within-participant (1^st^ day vs 2^nd^ day) variation in log-transformed 24-hour mean kitchen area NO_2_ concentration using a random intercept for the (two-day) sample. We treated post-intervention samples (3-, 6-, 12-month) as independent samples, and analyzed control (N = 76 paired samples) and LPG intervention (N = 84 paired samples) groups independently. Using the results from the mixed effects ANOVA, we calculated the intraclass correlation coefficient (ICC), which describes between-participant variance as a proportion of the total variance. We also calculated the coefficient of variation (CV) for 1^st^ and 2^nd^ day samples to assess the reproducibility of a one-day kitchen area NO_2_ sample when compared to the subsequent day.

#### 2.4.5. Analysis of variance of 1^st^ versus 2^nd^ post-intervention time points

We analyzed the reproducibility of collecting single versus multiple longitudinal NO_2_ samples by exploring within-participant versus between-participant variance of kitchen area samples taken months apart during the post-intervention period. We included in the analysis the first two valid samples from the post-intervention period for each participant. We used only the 1^st^ day 24-hour mean from each 48-hour sample to improve the comparison with results from the 1^st^ day vs 2^nd^ consecutive day variance analysis (Section 2.4.4.). We log-transformed 24-hour mean NO_2_ concentrations to comply with model assumptions of homoscedasticity of residuals. In our final model, we conducted a one-way mixed effects ANOVA with a random intercept for household, analyzing intervention and control groups independently and calculating the ICC for between-household variance. Additionally, we calculated the treatment group-specific coefficient of variation for 1^st^ and 2^nd^ post-intervention samples to quantify the reproducibility of kitchen area NO_2_ samples taken longitudinally throughout the post-intervention period of the trial. All analyses were performed using R (www.r-project.org).

## 3. Results

### 3.1 Participant characteristics

We sampled kitchen area NO_2_ concentrations using direct-reading monitors among 49 participants in the LPG intervention group and 47 participants in the control group (total N=96 participants). Due to battery failures, four participants (4% of N=100) did not have any post-intervention samples reaching the minimum duration (20 hours) and were excluded from the analysis. The mean age among all participants in the NO_2_ assessment was 48.2 years and 59% of participants had a primary school education or less (**Table 1**). Ninety-three percent of participants were in the lowest two quintiles of socio-economic status in Peru. Only 6% of intervention participants’ kitchens had a chimney, while 67% had an opening in the roof above the biomass cookstove and 27% had no specific cookstove ventilation. This differed somewhat from control participants, who had more chimneys (13%), fewer roof openings (38%), and more homes with no cookstove ventilation (49%). Typical kitchens among study participants had roofs of corrugated metal or natural fiber, walls of adobe or mud, and earth floors. Many kitchens had no windows (40%), while 44% of kitchens had one window and 17% of kitchens had two or more windows. Using the SUMs which monitored both LPG and biomass cookstoves continuously in all participants’ homes, we estimated that women in the LPG intervention group used their LPG stoves exclusively in 98% of monitored days.

**Table 1.** Characteristics of study participants and their kitchens in Puno, Peru.

|  | Intervention Homes | Control Homes | All Homes |
| --- | --- | --- | --- |
|  | N (%) or<br>Mean (SD) | N (%) or<br>Mean (SD) | N (%) or<br>Mean (SD) |
| Number of participants | 49 | 47 | 96 |
| Age in years | 49.3 (8.5) | 47.1 (11.9) | 48.2 (10.3) |
| Education |  |  |  |
| Primary or less | 31 (63) | 28 (60) | 59 (61) |
| Secondary | 18 (37) | 19 (40) | 37 (39) |
| National SES quintile |  |  |  |
| 1 (lowest) | 24 (49) | 28 (60) | 52 (54) |
| 2 | 21 (43) | 16 (34) | 37 (39) |
| 3 | 4 (8) | 3 (6) | 7 (7) |
| 4 | 0 (0) | 0 (0) | 0 (0) |
| 5 (highest) | 0 (0) | 0 (0) | 0 (0) |
| Cookstove ventilation |  |  |  |
| Chimney | 3 (6) | 6 (13) | 9 (9) |
| Roof opening | 33 (67) | 18 (38) | 51 (53) |
| No cookstove ventilation | 13 (27) | 23 (49) | 36 (38) |
| Roof type |  |  |  |
| Corrugated metal | 21 (43) | 16 (34) | 37 (39) |
| Natural fiber (thatch) | 27 (55) | 30 (64) | 57 (59) |
| Other | 1 (2) | 1 (2) | 2 (2) |
| Wall type |  |  |  |
| Adobe/mud with plaster | 14 (29) | 13 (28) | 27 (28) |
| Adobe/mud without plaster | 31 (63) | 32 (68) | 63 (66) |
| Other | 4 (8) | 2 (4) | 6 (6) |
| Floor type |  |  |  |
| Dirt | 45 (92) | 44 (94) | 89 (93) |
| Cement | 4 (8) | 3 (6) | 7 (7) |
| Kitchen windows (#) |  |  |  |
| 0 | 19 (39) | 19 (40) | 38 (40) |
| 1 | 24 (49) | 18 (38) | 42 (44) |
| 2 + | 6 (12) | 10 (21) | 16 (17) |
| Kitchen doors/entryways (#) |  |  |  |
| 1 | 49 (100) | 47 (100) | 96 (100) |

### 3.2. Post-intervention kitchen area nitrogen dioxide concentrations

During the post-intervention period and using direct-reading monitors, we successfully collected 367 24-hour mean kitchen area concentrations from 207 samples (20-48 hours duration) representing a total of 96 unique households from the intervention and control groups. We observed a geometric mean (GM) 24-hour kitchen area NO_2_ concentration of 43 ppb (geometric standard deviation [GSD] 1.7) in the LPG intervention group during the post-intervention period, 30% higher than the WHO indoor annual guideline of 33 ppb (**Table 2**). Sixty-nine percent of LPG intervention kitchen samples had 24-hour mean concentrations that exceeded the WHO indoor annual guideline. In control kitchens, the GM 24-hour kitchen area concentration during the post-intervention period was 77 ppb (GSD 2.0). Kitchen area NO_2_ concentrations exceeded the WHO indoor hourly guideline for a mean of 1.3 hours per day in intervention households and 2.5 hours per day in control households. We observed a GM kitchen concentration of 91 ppb (GSD 2.1) during LPG cooking events in the intervention group, compared to a GM concentration of 33 ppb (GSD 1.8) outside of LPG cooking events (though the mean concentration outside of cooking events includes time directly after cooking events ended, when NO_2_ concentrations likely remained elevated before decaying to background levels). In control households, GM kitchen area concentrations were 296 ppb (GSD 2.8) during biomass cooking events and 39 ppb (GSD 2.0) outside of recorded cooking events. A subset of participants received additional kitchen area sampling of 48-hour time-weighted average concentration via passive samplers. Among 37 post-intervention samples from 16 unique participants in the LPG intervention group, we observed a GM 48-hour mean kitchen area concentration of 29 ppb (GSD 2.2). In the control group, we observed a GM 48-hour kitchen area mean of 99 ppb (GSD 4.3) in 21 post-intervention samples from 9 unique participants (**Table 2**).

**Table 2.** Nitrogen dioxide kitchen concentrations and personal exposures among women in the post-intervention period of a biomass-to-LPG cookstove intervention trial in Puno, Peru.

|  | LPG Intervention |  |  |  |  |  |  | Biomass Control |  |  |  |  |  |  |
| --- | --- | --- | --- | --- | --- | --- | --- | --- | --- | --- | --- | --- | --- | --- |
|  | N | Mean | SD | GM | GSD | Median | IQR | N | Mean | SD | GM | GSD | Median | IQR |
| Kitchen area: direct-reading |  |  |  |  |  |  |  |  |  |  |  |  |  |  |
| Maximum 1-hr rolling means (ppb) | 179 | 178 | 126 | 138 | 2.1 | 149 | 168 | 188 | 748 | 697 | 450 | 3.1 | 543 | 840 |
| 24-hr means (ppb) | 179 | 49 | 26 | 43 | 1.7 | 42 | 29 | 188 | 96 | 65 | 77 | 2.0 | 81 | 79 |
| Daily hours > 163 ppb | 179 | 1.3 | 1.6 | - | - | 0.6 | 1.8 | 188 | 2.5 | 2.1 | - | - | 1.8 | 2.4 |
| Means during cooking (ppb) | 102* | 114 | 72 | 91 | 2.1 | 91 | 105 | 89* | 455 | 397 | 296 | 2.8 | 377 | 450 |
| Kitchen area: passive badge |  |  |  |  |  |  |  |  |  |  |  |  |  |  |
| 48-hr means (ppb) | 37 | 38 | 29 | 29 | 2.2 | 31 | 32 | 21 | 185 | 162 | 99 | 4.3 | 129 | 178 |
| Personal exposure: passive badge |  |  |  |  |  |  |  |  |  |  |  |  |  |  |
| 48-hr means (ppb) | 35 | 8 | 11 | 5 | 2.4 | 4 | 5 | 21 | 23 | 24 | 16 | 2.3 | 17 | 18 |

\*Concentrations during cooking events were calculated over the entire available sample duration, not divided into multiple 24-hour averages

We observed acute spikes in NO_2_ kitchen area concentrations during common cooking times among participants in both the intervention and control groups. We present these data as a bar plot of kitchen area concentrations throughout each minute of the calendar day (**Figure 2)** from all post-intervention samples. Dark blue indicates the proportion of households with kitchen area NO_2_ concentrations <= 32 ppb at a given time of day, with increasingly higher concentrations represented by other colors as described in the legend. A substantial proportion of kitchens in the LPG intervention group (**Figure 2**, top panel) experience NO_2_ concentrations exceeding WHO indoor guidelines (annual 33 ppb, hourly 163 ppb) during common cooking times (05:00-09:00 and 18:00-20:00 hours). For example, at approximately 08:00 hours, NO_2_ concentrations were ≥ 250 ppb in 15% of households (red color), ≥ 163 ppb (the WHO indoor hourly guideline) in 25% of households (red and orange colors), and ≥ 66 ppb in 55% of households (red, orange, and yellow colors). In the corresponding figure of NO_2_ concentrations in biomass cookstoves (**Figure 2**, bottom panel), concentrations are elevated during the same common cooking hours, but peaks are at higher concentrations in biomass homes than in LPG homes. The GM highest hourly concentration during each 24-hour sample was 138 ppb (GSD 2.1) in LPG intervention homes and 450 ppb (GSD 3.1) in biomass control households (**Table 2)**. We present the distribution of highest hourly means in the intervention and control groups as a modified empirical distribution function plot (**Figure 3)**, with the WHO indoor hourly guideline as a reference. The X-axis represents NO_2_ concentration and the Y-axis represents the percent of 24-hour samples with a maximum hourly-average concentration less than the corresponding concentration. During the intervention period 47% of 24-hour samples in the LPG intervention group and 81% of 24-hour samples in the biomass control group had hourly means exceeding the WHO indoor hourly guideline.

**Figure 2.**
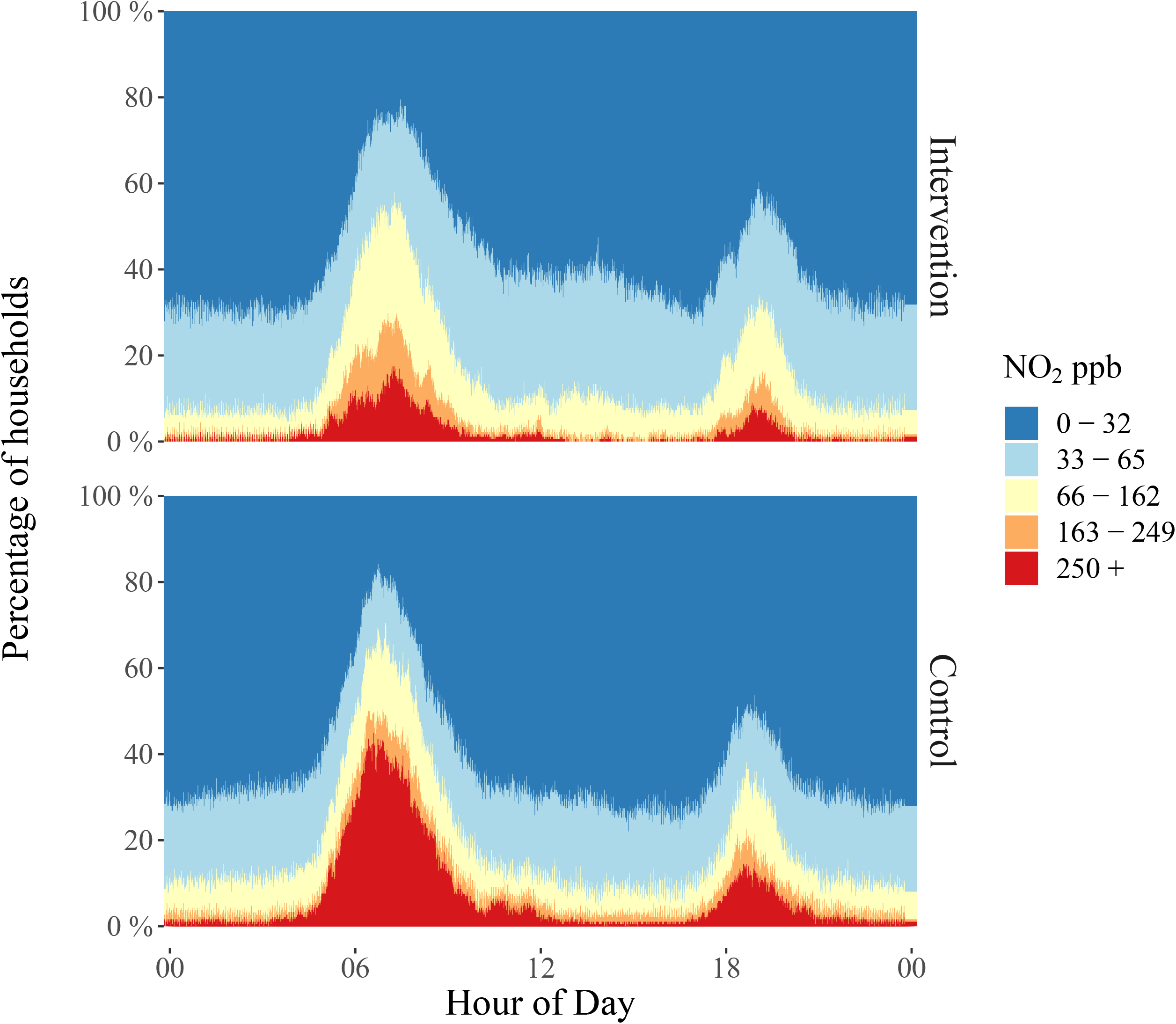
Prevalence of kitchen area NO_2_ concentrations by calendar minute in 179 24-hour samples from 49 houses in the intervention group and 188 24-hour samples from 47 houses in the control group of a biomass-to-LPG cleaner-cooking trial in Puno, Peru.

**Figure 3.**
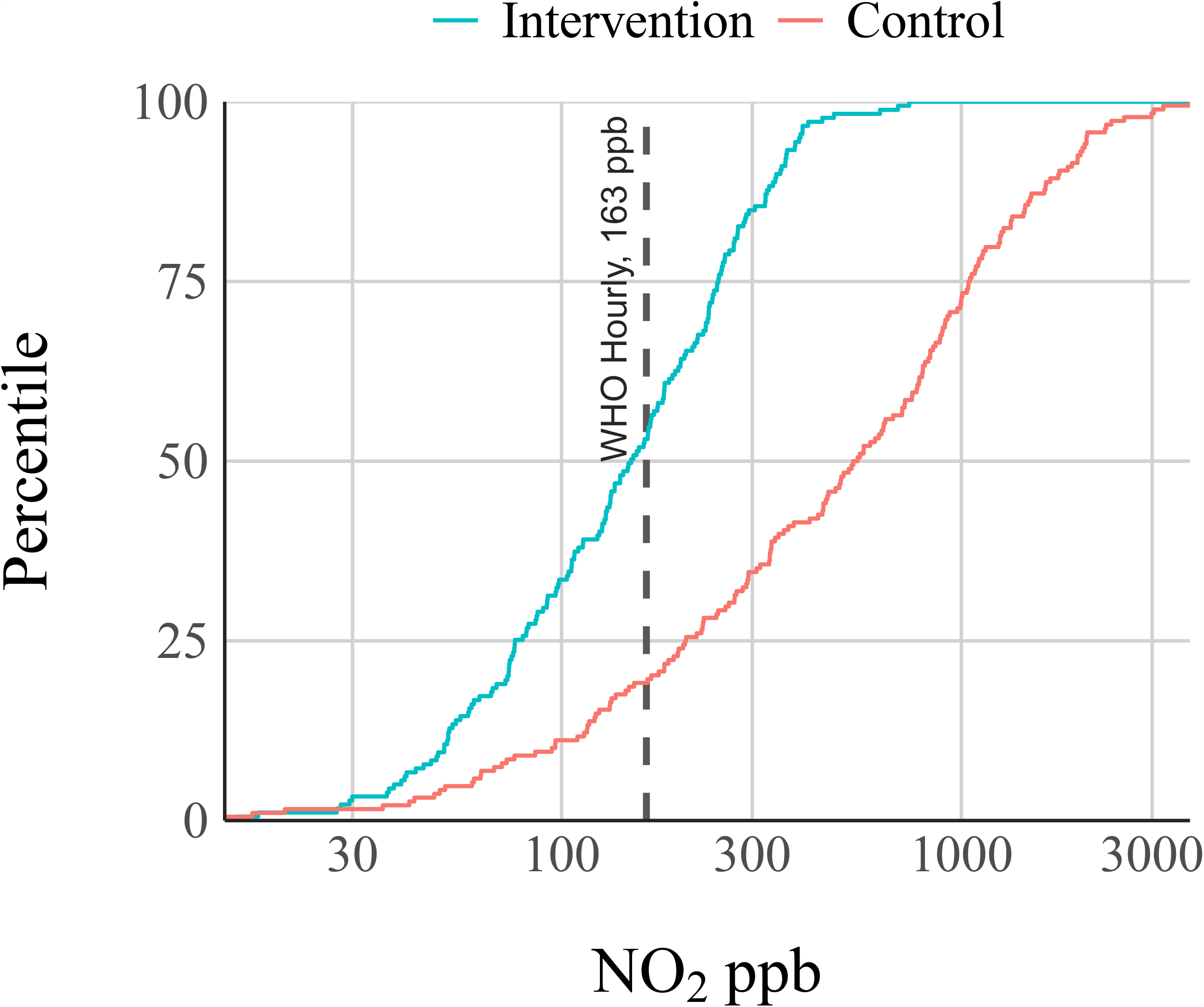
Cumulative distributions of the highest hourly mean NO_2_ concentrations in 367 24-hour samples of 96 kitchen areas, comparing intervention and control groups during the follow-up period of a biomass-to-LPG intervention trial in Puno, Peru.

### 3.3. Personal exposure to nitrogen dioxide

Among 35 samples from 16 unique participants in the LPG intervention group, we observed a 48-hour mean NO_2_ personal exposure of 8 ppb (SD 11 ppb) with a GM of 5 ppb (GSD 2.4). We observed a mean of 23 ppb (SD 24 ppb) and a geometric mean of 16 ppb (geometric SD 2.3 ppb) 48-hour personal exposure among 21 samples from 9 participants in the control group. Three percent (N = 1 of 35) of personal exposure samples from women in the LPG intervention group and 19% (N = 4 of 21) of personal exposure samples in the control group had 48-hour time-integrated personal exposures in excess of the WHO indoor annual guideline of 33 ppb.

### 3.4. Longitudinal effect of LPG intervention on NO_2_ exposures

In **Figure 4**, kitchen area 24-hour means are presented from baseline through the end of the post-intervention period, with lines indicating treatment group means at each time point, points representing individual 24-hour mean concentrations, and the WHO indoor annual guideline added for reference. Using a one-way ANOVA, we found no evidence of longitudinal differences in group means across post-intervention time points in either 179 24-hour means from 49 participants in the LPG intervention group (p-value = 0.09) or 188 24-hour means from 47 participants in the control group (p-value = 0.99).

**Figure 4.**
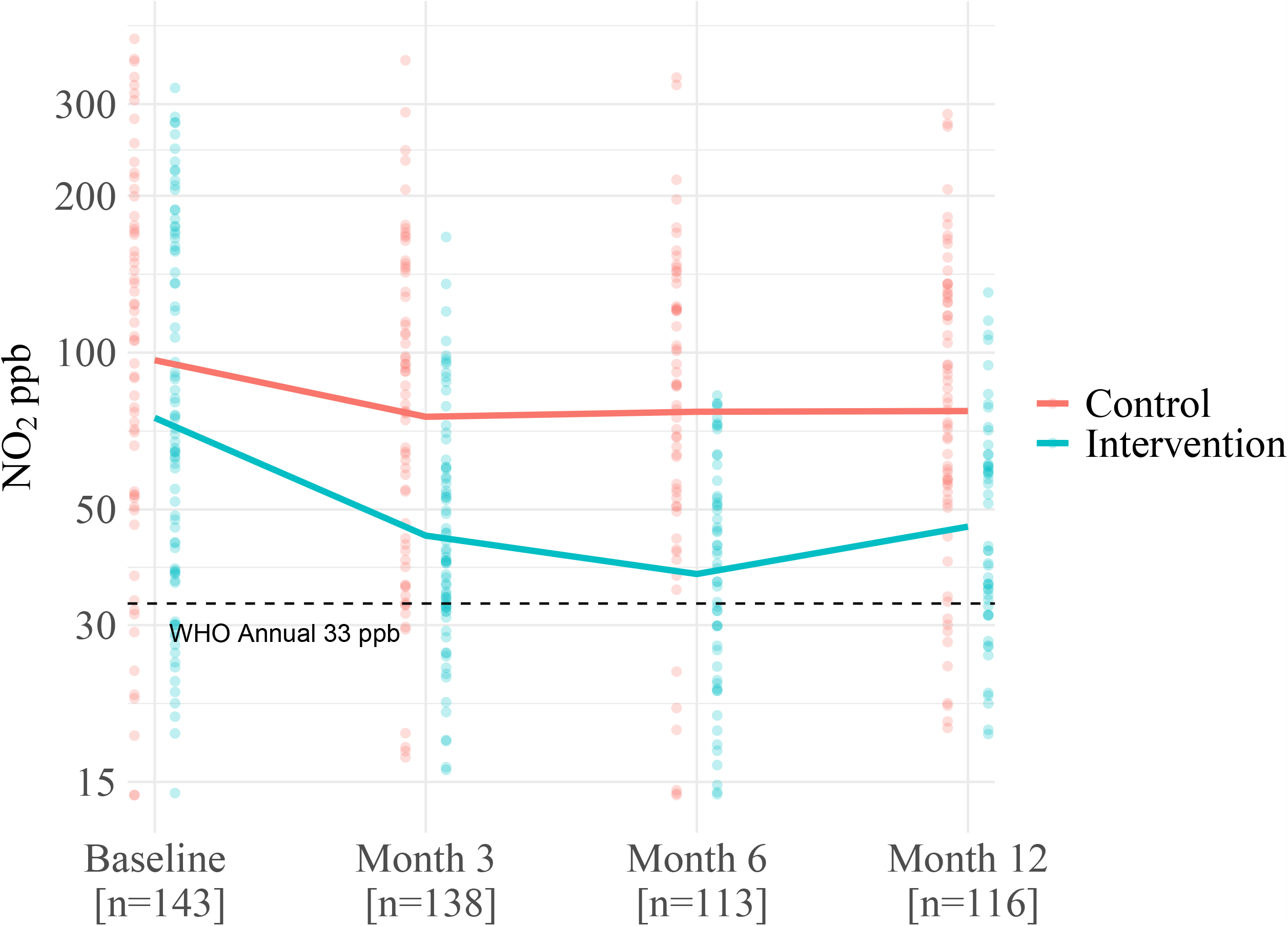
Longitudinal changes in kitchen area 24-hour mean NO_2_ concentrations among intervention and control groups in an LPG intervention trial. Lines indicate mean kitchen area NO_2_ concentrations at each time point for the intervention and control groups. Points represent NO_2_ 24-hour mean concentration from 367 samples in 96 unique households. The Y-axis representing NO_2_ ppb is log-scaled. Altitude- and temperature-adjusted WHO indoor air quality guideline for annual mean NO_2_ (33 ppb) presented as a reference.

Because baseline kitchen area concentrations were lower in the LPG intervention group, we used linear regression to estimate the effect of treatment group on post-intervention kitchen area NO_2_, adjusting for baseline concentration (Section 2.4.3.). We estimate that among 79 participants with baseline and post-intervention samples, being in the LPG intervention group was associated with a 45 ppb lower (95% CI −59 to −31) post-intervention daily mean kitchen area concentration when compared to the control group.

### 3.5. Between- and within-variation among 1^st^ versus 2^nd^ consecutive sampling days

We examined between-participant versus within-participant variance among kitchen area 24-hour means on the 1^st^ versus 2^nd^ consecutive days of sampling (Section 2.4.4.). In both the LPG intervention and control groups, we found greater variance between households than within households, however the reproducibility of sampling within a household on consecutive days was somewhat poor. Within 76 paired samples (1^st^ and 2^nd^ consecutive days) in the LPG intervention group we observed an intraclass correlation coefficient (ICC) of 0.68, indicating that 68% of the total variance was between households while 32% of total variance was within households (**Table 3**). Similarly, we found that 73% of variance was between households (ICC 0.73) with a CV of 35% in 84 paired samples of the biomass control group.

**Table 3.**
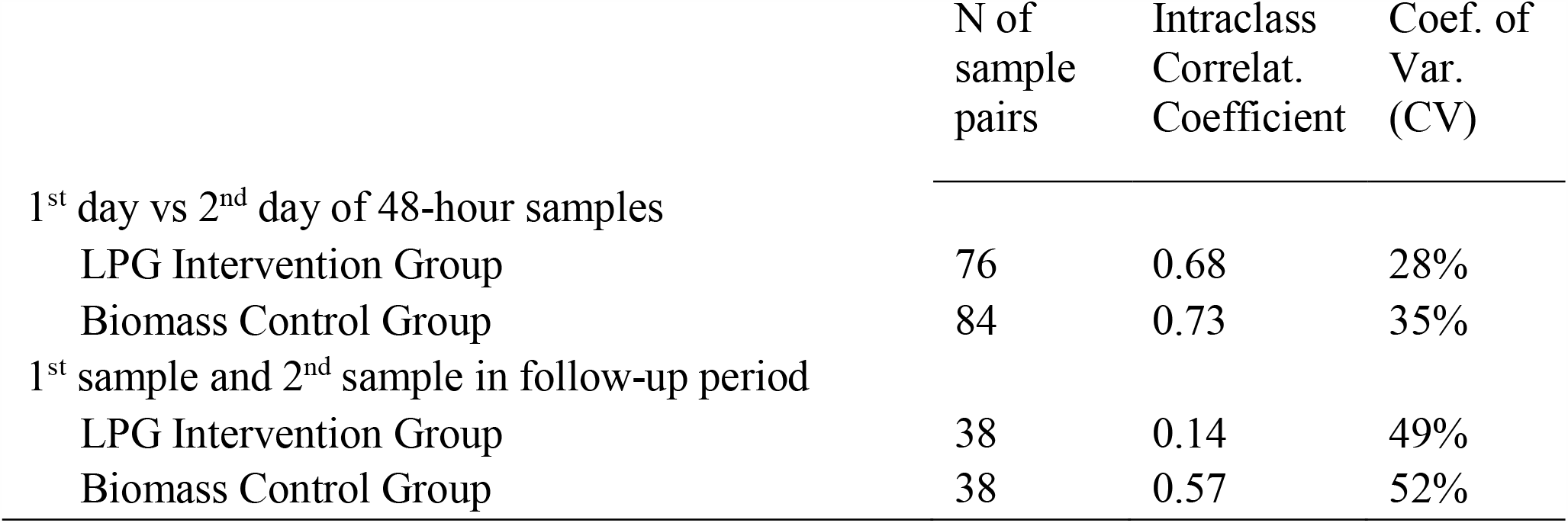
Analysis of variance of kitchen area NO2 concentrations between and within 1) consecutive sample days and 2) repeated samples throughout the study follow-up period of an LPG cookstove intervention trial in Puno, Peru.

|  | N of<br>sample<br>pairs | Intraclass<br>Correlat.<br>Coefficient | Coef. of<br>Var.<br>(CV) |
| --- | --- | --- | --- |
| 1<sup>st</sup> day vs 2<sup>nd</sup> day of 48-hour samples |  |  |  |
| LPG Intervention Group | 76 | 0.68 | 28% |
| Biomass Control Group | 84 | 0.73 | 35% |
| 1<sup>st</sup> sample and 2<sup>nd</sup> sample in follow-up period |  |  |  |
| LPG Intervention Group | 38 | 0.14 | 49% |
| Biomass Control Group | 38 | 0.57 | 52% |

### 3.6. Between- and within- variation among 1^st^ versus 2^nd^ post-intervention time points

We also compared kitchen area NO_2_ concentrations from longitudinal samples taken months apart during the post-intervention period (Section 2.4.5.). In the LPG intervention group, we observed an ICC of 0.14 among 38 sample pairs (1^st^ and 2^nd^ available post-intervention time points), indicating more variance within a given household across time (86% of total variance) than between different households (14% of total variance). Within the same group we estimated a CV of 49%, suggesting poor reproducibility within participants over time. In the control group of 38 sample pairs, the ICC was 0.57 with a CV of 52%, suggesting a more equal balance of between/within variance but similarly poor reproducibility across the post-intervention period.

## 4. Discussion

This study is the first study that the authors are aware of to measure kitchen area concentrations of NO_2_ at high-temporal resolution or personal exposure to NO_2_ from LPG stoves in an LMIC field setting. We observed substantial reductions in kitchen area concentration and personal exposure to NO_2_ in a biomass-to-LPG intervention. While lower than biomass control households, in the LPG intervention group, we observed large concentrations spikes of kitchen area NO_2_ concentrations during common cooking times. In the LPG intervention group, 69% of 24-hour samples exceeded the WHO indoor annual guideline and 47% of samples exceeded the WHO indoor hourly guideline. Participants in the LPG intervention group experienced a mean of 1.3 hours per day (SD 1.6 hours) of kitchen area NO_2_ concentrations above the WHO indoor hourly guideline. GM 24-hour average kitchen area concentrations were 1.3 times higher than the WHO indoor annual guideline (163 ppb) in the LPG intervention group and 2.3 times higher in the biomass control group. However, GM 48-hour mean personal exposure was well below WHO indoor annual guidelines in the LPG intervention group.

Among homes using LPG stoves, we observed an arithmetic mean 24-hour kitchen area NO_2_ concentration of 49 ppb (SD 26 ppb) using direct-reading monitors and 38 ppb (SD 29 ppb) in a subset of homes using passive samplers. These values are similar to a arithmetic mean of 38 ppb NO_2_ reported by Padhi et al. among 24-hour samples of kitchens with LPG stoves in India.^20^ While assessments of NO_2_ exposures from LPG stoves in LMIC settings are sparse, a few other studies have reported NO_2_ concentrations in kitchens with natural gas or non-specific “gas stoves”. A study of kitchens with gas stoves in Bangladesh reported a 24-hour geometric mean kitchen area NO_2_ concentration of 84 ppb,^17^ though the specific type of gas fuel (i.e. natural gas, LPG, other) was not reported. Colbeck et al. observed 1-week mean NO_2_ concentrations in kitchens with natural gas stoves in Pakistan of 129 ppb in the winter when windows are kept closed and 43 ppb in the summer when windows are open,^18^ suggesting that ventilation may be an important and actionable predictor of indoor NO_2_ concentrations in homes with gas stoves. This was corroborated on a smaller magnitude among LPG intervention participants in our study, in which mean kitchen area concentrations were 52 ppb (SD 29 ppb, N = 53 24-hour samples) in winter and 45 ppb (SD 20 ppb, N = 27 24-hour samples) in summer.

The relative NO_2_ emissions of LPG stoves vs natural gas stoves in LMIC field settings is poorly understood, and other factors which have major impacts on area concentrations, including stove burner design, individual cooking behaviors, and kitchen size and ventilation are rarely available in the current literature. In a seminal review of NO_2_ exposures from gas stoves in HICs, where overall stove quality is potentially higher than in many LMIC settings, use of a gas stove increased mean indoor NO_2_ by 15 ppb compared to homes with electric stoves. In this review, an equivalent 15 ppb increase in indoor area NO_2_ concentration corresponded with an odds ratio of 1.18 for lower respiratory tract illnesses in children.^53^ In homes with LPG stoves in Puno, we observed a GM 24-hour kitchen area NO_2_ concentration of 43 ppb in the LPG intervention group, 10 ppb higher than the WHO indoor annual guideline of 33 ppb. We also observed concentration spikes that commonly exceeded 500 ppb and a mean maximum hourly mean kitchen area concentration of 178 ppb (WHO indoor hourly guideline: 163 ppb). Concentration spikes on this order of magnitude have been reported previously in homes with gas appliances in HICs. For example, a field study of children in Australian homes with gas stoves found 1-hour mean personal exposures of greater than 200 ppb during periods of gas stove use.^67^

We assessed the between-participant vs within-participant variance of measuring kitchen area NO_2_ on two consecutive days during the post-intervention period. We found greater between-participant variance than within-participant variance, suggesting that limited sampling resources may be more efficiently directed towards sampling a larger number of participants for 24-hours than fewer participants for 48-hours. However, 24-hour kitchen area NO_2_ samples had poor reproducibility on consecutive days, and the limitations of a 24-hour kitchen area samples should be considered when designing studies which are focused on individual-level health outcomes, where personal exposure levels are more relevant.

We also analyzed the between-participant vs within-participant variance of kitchen area NO_2_ measurements taken months apart during the post-intervention period. Compared to the analysis of samples on subsequent days, we found more within-participant variability among samples taken months apart, which may be related to seasonality. Within-participant variability was similar between the LPG intervention and biomass control groups, but between-participant variability was substantially lower in the LPG group (16% of total variance). This could be explained by more similarity in emissions from LPG stoves than biomass stoves due to standardization of the stoves and fuel, which were provided to participants in the intervention trial. In contrast, biomass stoves are often homemade and can use a variety of biomass fuel types. It may be that given a standardized LPG intervention, a relatively small number of participants are needed to reasonably assess NO_2_ kitchen area concentrations in the group longitudinally, though likely only in settings where other emissions-related factors such as kitchen ventilation are also consistent. It is worth noting that in this intervention trial, we observed 98% exclusive adoption of LPG stoves and consistency in NO_2_ concentrations longitudinally across post-intervention samples, and it is highly unlikely that the observed levels of NO_2_ are due to continued use of biomass stoves in the LPG intervention arm.

This study is strengthened by its use of direct-reading monitors, which allowed us to characterize concentration spikes associated with LPG cooking and compare kitchen area concentrations with WHO indoor hourly air quality guidelines, which have not been previously reported. By deploying stove use monitors, we were also able to co-monitor stove use and kitchen area NO_2_ concentration to estimate concentrations during cooking events and the duration of time per day spent above WHO indoor guidelines. We also measured 48-hour mean personal exposure to NO_2_ among a subsample of LPG and biomass users, which is a novel contribution to the field. This study was further strengthened by the use of longitudinal measurements throughout a cleaner-cooking intervention with a one-year follow-up period. This study is limited by a lack of measurements of hourly or peak personal exposure to NO_2_, due to the burden of asking participants to carry NO_2_ direct-reading monitors. Based on the observed high concentration spikes of kitchen area NO_2_ concentrations during cooking and studies in HICs, we believe the greatest risk of exposure to NO_2_ for people who use LPG stoves are concentration spikes as opposed to mean NO_2_ concentrations. While many women in our setting may not spend the entire duration of a cooking event in the kitchen area, the peak personal exposures of women in our setting may in fact be comparable to the concentration spikes observed in the kitchen areas when they are actively cooking. However, 48-hour mean personal exposures to NO_2_ were well below WHO indoor annual guidelines for most participants in the LPG intervention group. Future research is warranted to characterize personal exposure to LPG stove-related NO_2_ concentration spikes, assess personal exposure among children who are especially vulnerable to NO_2_ exposure, and to compare NO_2_ exposures in households with LPG stoves to households with electric stoves in LMICs.

## 5. Conclusions

In a biomass-to-LPG intervention trial in the Peruvian Andes, we observed substantially lower NO_2_ kitchen area concentrations and personal exposures among participants in the LPG intervention. However, within LPG intervention households, 69% of 24-hour samples of kitchen area concentration exceeded WHO indoor annual guidelines and 47% of samples exceeded WHO indoor hourly guidelines. Among a subsample of participants, GM 48-hour personal exposure was well below WHO indoor annual guidelines in the LPG intervention group.

While measurements of NO_2_ concentrations from LPG stoves are sparse in LMICs, these results are not unexpected given previous assessments of NO_2_ in kitchens with gas stoves in LMICs and the growing body of literature on the health impacts of NO_2_ exposures from gas stoves in HICs. As the global community considers the promotion of LPG and other gas stoves as cleaner-burning alternatives to biomass based on reductions in PM_2.5_ and CO, exposures to NO_2_ emitted by LPG stoves may persist at levels that pose a risk to health. In settings where LPG stoves are currently being used or use of electric stoves is still far off, the ability of actionable factors such as ventilation and stove design to mitigate NO_2_ exposures should be explored further and incorporated into LPG promotion campaigns.

## Data Availability

Available upon reasonable request.

## Acknowledgements

Financial support for the CHAP trial was received from the Global Environmental and Occupational Health, Fogarty International Center, United States National Institutes of Health (U01TW010107 and U2RTW010114); the Clean Cooking Alliance of the United Nations Foundation (UNF 16-810), the Johns Hopkins Center for Global Health, and the COPD Discovery Fund of Johns Hopkins University. The Center for Global Non-Communicable Disease Research and Training field site in Puno, Peru, also received generous support from Mr. William and Bonnie Clarke III.

JLK and KNW were supported by the NIH Fogarty International Center, NINDS, NIMH, NHBLI and NIEHS under NIH Research Training Grant # D43 TW009340 and the Johns Hopkins Center for Global Health. JLK was also supported by the Ruth L. Kirschstein Institutional National Research Service Award (5T32ES007141-33) funded by the NIH/NIEHS. KNW was also supported by the National Heart, Lung, and Blood Institute of the National Institutes of Health under Award Number T32HL007534. MFDR was further supported by the Global Environmental and Occupational Health (GEOHealth), Fogarty International Center, and by the David Leslie Swift Fund of the Bloomberg School of Public Health, Johns Hopkins University. The content is solely the responsibility of the authors and does not necessarily represent the official views of the National Institutes of Health.

## Appendix Statistical methods for analysis of stove temperature monitors

We developed separate empirical algorithms to predict LPG and biomass cookstove use with recorded stove temperatures. To identify LPG stove use, we considered an LPG cooking event to begin at time *t* when the 20-minute rolling mean temperature at time *t + 5 minutes* was at least 10 % greater than at *t – 5 minutes* (depicted in **Appendix Figure 1**). A cooking event stopped when the 20-minute rolling mean temperature dropped 3°C below the maximum 20-minute rolling mean temperature in the cooking event. For biomass cookstoves, we considered a cookstove usage event to begin at time *t* when the 30-minute rolling mean temperature at time *t + 30 minutes* was 2°C greater than at *t*. A cooking event stopped when the 30-minute rolling mean temperature dropped 2°C below the maximum 30-minute rolling mean temperature in the cooking event. For both types of cookstove, we made *a priori* assumptions based on formative research that multiple cooking events within a 60-minute period were considered one cooking event, an individual cooking event cannot last more than four hours for an LPG stove or six hours for a biomass cookstove, and the rolling mean must exceed 20°C at some point during a cooking event. To assess the validity of the SUMs algorithms, an independent researcher not involved in the creation of the algorithm manually evaluated a 5-day random sample of SUMs data from each stove in each household in CHAP (N=180 households). Manual observations and algorithm estimates were in agreement on whether stove use had occurred in 95% of 787 days of monitored biomass cookstoves and in 99.7% of 762 days of monitored LPG stoves.

**Appendix Figure 1.**
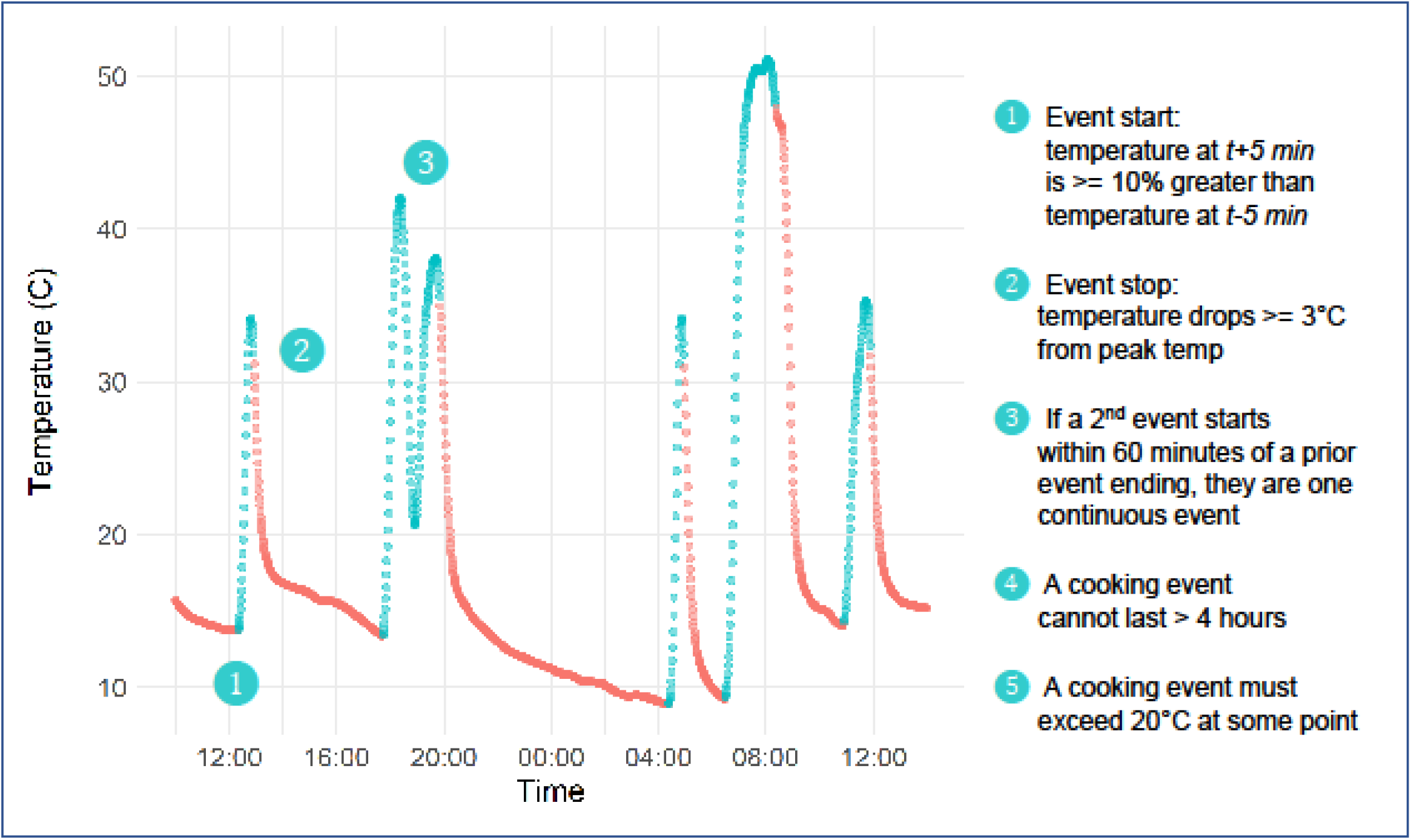
Empirical algorithm for identifying LPG stove use from stove temperature logged throughout the duration of the study at one-minute intervals. A similar algorithm exists for biomass cookstoves (not shown).

